# From Isolation to Coordination: How Can Telemedicine Help Combat the COVID-19 Outbreak?

**DOI:** 10.1101/2020.02.20.20025957

**Authors:** Yunkai Zhai, Yichuan Wang, Minhao Zhang, Jody Hoffer Gittell, Shuai Jiang, Baozhan Chen, Fangfang Cui, Xianying He, Jie Zhao, Xiaojun Wang

## Abstract

The rapid spread of Coronavirus disease 2019 (COVID-19) presents China with a critical challenge. As normal capacity of the Chinese hospitals is exceeded, healthcare professionals struggling to manage this unprecedented crisis face the difficult question of how best to coordinate the medical resources used in highly separated locations. Responding rapidly to this crisis, the National Telemedicine Center of China (NTCC), located in Zhengzhou, Henan Province, has established the Emergency Telemedicine Consultation System (ETCS), a telemedicine-enabled outbreak alert and response network. ETCS is built upon a doctor-to-doctor (D2D) approach, in which health services can be accessed remotely through terminals across hospitals. The system architecture of ETCS comprises three major architectural layers: (1) telemedicine service platform layer, (2) telemedicine cloud layer, and (3) telemedicine service application layer. Our ETCS has demonstrated substantial benefits in terms of the effectiveness of consultations and remote patient monitoring, multidisciplinary care, and prevention education and training.

---

The rapid spread of Coronavirus disease 2019 (COVID-19) presents China with a critical challenge. As normal capacity of the Chinese hospitals is exceeded, healthcare professionals struggling to manage this unprecedented crisis face the difficult question of how best to coordinate the medical resources used in highly separated locations. Many cities in China have been imposing a lockdown, due to the high risk of infection and the characteristic of human-to-human transmission^1^. Not only have patients been marginalized, but many clinicians working in the regional hospitals have limited access to the specialist consultations and treatment guidelines they need from provincial-level hospitals to manage pneumonia cases caused by COVID-19. As long as the crisis continues, simply relying on the traditional communicative practices, such as physician office visit or face-to-face consultations within the health professional network, could pose significant costs and health concerns.

Telemedicine has been acknowledged as a breakthrough technology in combating epidemics^2^. Combining the functions of online conversation and real-time clinical data exchange, telemedicine can provide technical support to the emerging need for workflow digitalization. When facing the rapid spread of an epidemic, the ability to deliver clinical care in a timely manner requires effective relational coordination mechanisms amongst government authorities, hospitals, and patients^3^. This raises the question: ***How can telemedicine systems operate in a coordinated manner to deliver effective care to patients with COVID-19 and to combat the crisis outbreak?***

Responding rapidly to this crisis, the National Telemedicine Center of China (NTCC), located in Zhengzhou, Henan Province, has established the Emergency Telemedicine Consultation System (ETCS), a telemedicine-enabled outbreak alert and response network. Implementation of the telemedicine system is supported by grants from the Novel Coronavirus Pneumonia Prevention and Control Headquarters (NCPPCH) and the Finance Department of Henan Province. A subsidy of 10 million Chinese yuan has been allocated to the construction and operation of ETCS. Collaborating with China Mobile and Huawei Technologies Co., Ltd., on January 29, 2020 the NTCC sent 18 workgroups to isolation wards to set up telemedicine networks and equipment. The total round-trip distance we travelled was about 9,320 miles, via box trucks, while the approximate round-trip distance to the farthest hospital from the NTCC headquarters in Zhengzhou was 743 miles. Based on our previous experience of building 5G networks and smart medical terminals, 126 network hospitals were successfully connected to the NTCC on ETCS within 82 hours.

ETCS is built upon a doctor-to-doctor (D2D) approach, in which health services can be accessed remotely through terminals across hospitals. The system architecture of ETCS comprises three major architectural layers: (1) telemedicine service platform layer, (2) telemedicine cloud layer, and (3) telemedicine service application layer, as depicted in Figure 1. ***The telemedicine service platform layer*** enables a specialist treatment team of provincial specialists to conquer distance and provide access to clinicians working in the regional hospitals. It provides clinicians and patients with immediate diagnosis and consultations regarding COVID-19, wireless remote patient monitoring, remote multiple disciplinary care, and telehealth for education and training, utilizing interactive live video conferencing. The specialist treatment team members work closely with the NCPPCH and Health Commission of Henan Province to prevent and control the spread of COVID-19. ***The telemedicine cloud layer*** allows clinicians to capture, store and process patient medical records, and to achieve real-time data exchange. In addition, prevention and treatment guidelines, and guidance on drug use and management of coronavirus patients can be accessed via the telemedicine cloud. ***The telemedicine service application layer*** involves 2 provincial-level hospitals, 18 municipal hospitals and 106 county-level hospitals, which can obtain consultations from the specialist treatment team. This logical structure supports a system of telemedicine clinical management to combat the COVID-19 outbreak.

**Figure 1.**
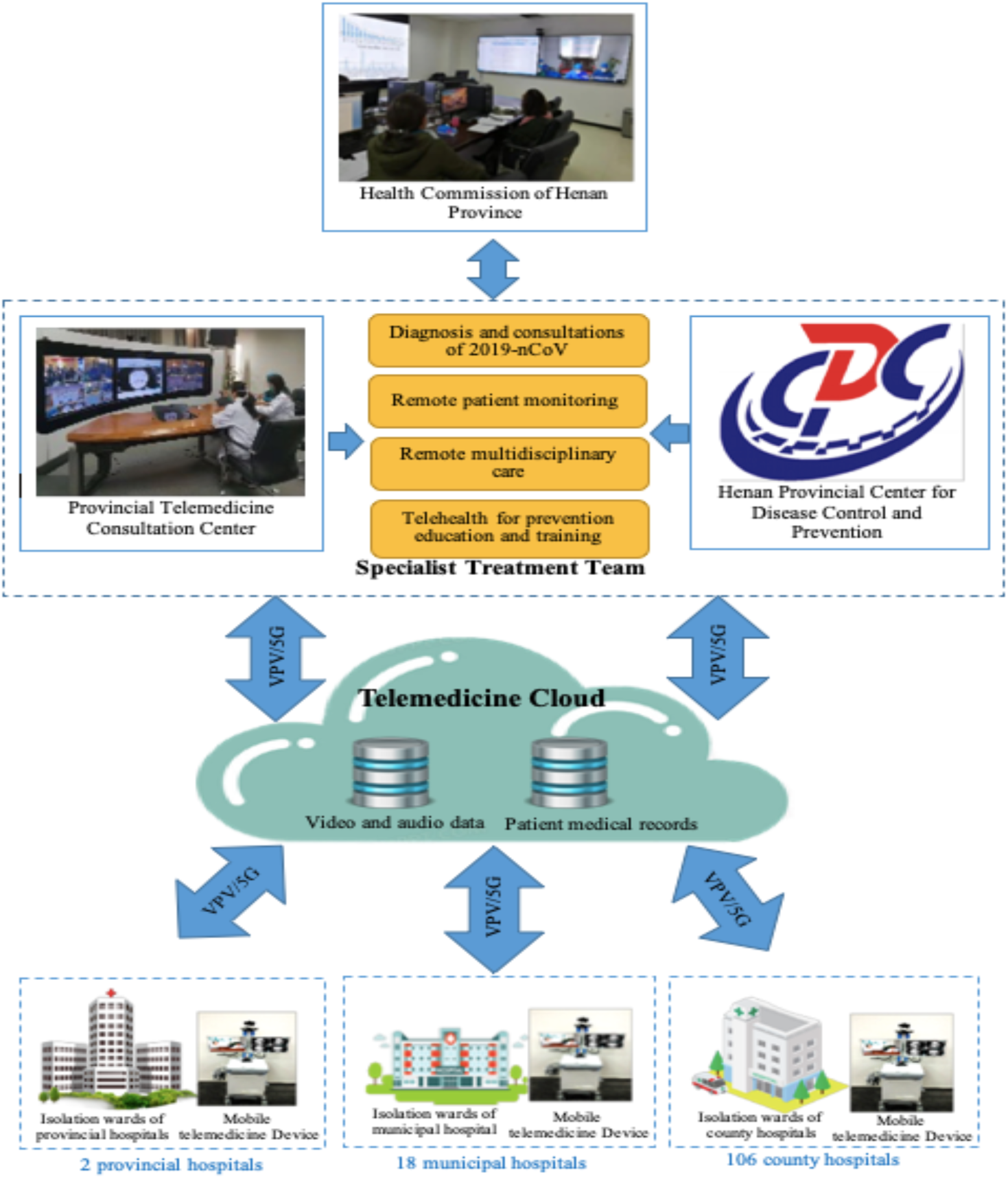
Architecture of the Emergency Telemedicine Consultation System.

**Figure 2.**
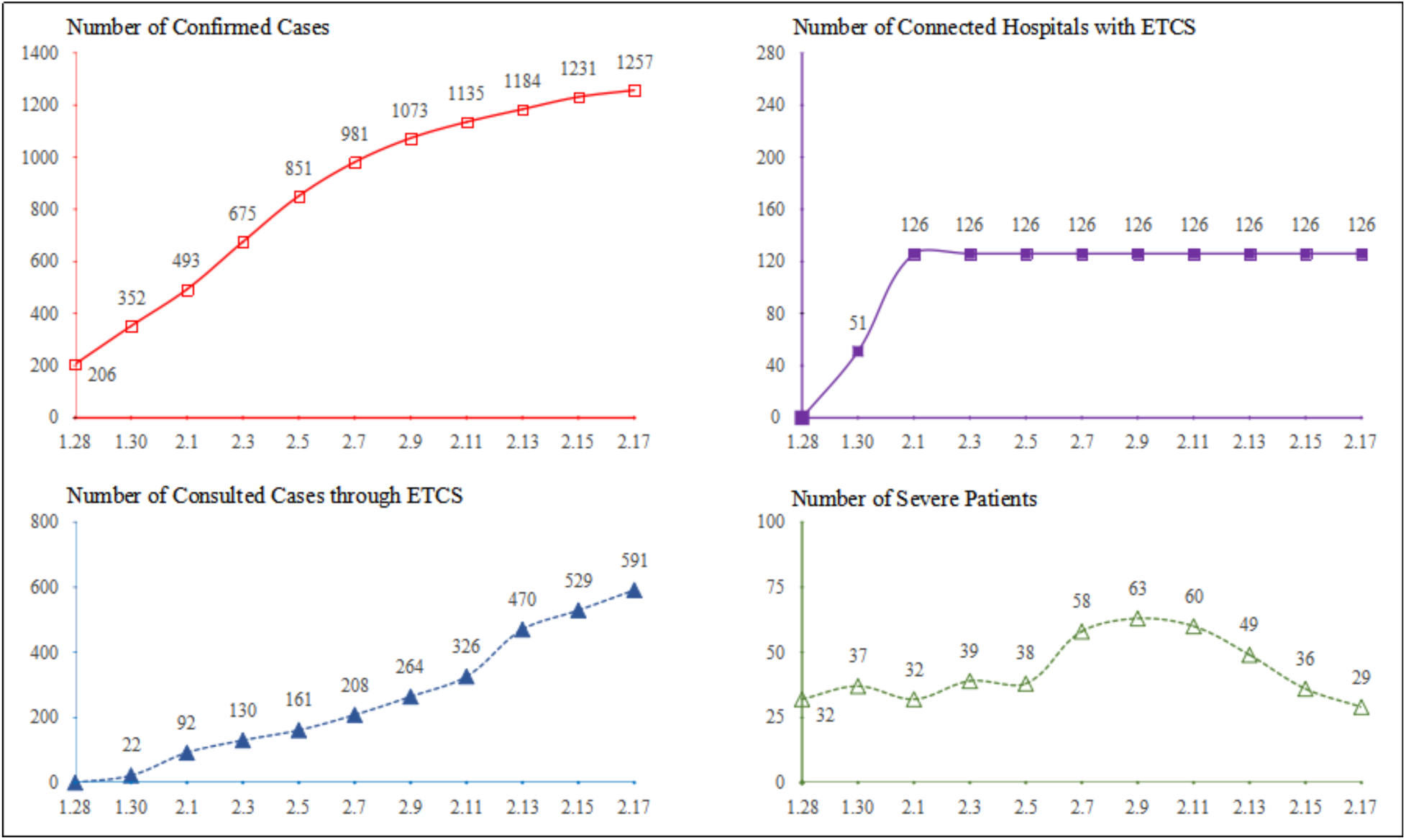
System performance of ETCS during the COVID-19 outbreak presented by the number of confirmed cases, the number of connected hospitals with ETCS, the number of consulted cases through ETCS, and the number of severe patients reported through ETCS Notes: 1.28 and 2.17 in the chart refer to January 28, 2020 and February 17, 2020 respectively. The number of consulted cases through ETCS refers to the number of consultations provided by the provincial specialist treatment team to the municipal and county hospitals, excluding the consultations provided by the municipal hospitals to the county hospitals.

Our ETCS has demonstrated substantial benefits in terms of the effectiveness of consultations and remote patient monitoring, multidisciplinary care, and prevention education and training. ***First***, 63 severe cases and 591 patients with mild and moderate respiratory infections received telemedicine consultations through the ETCS between January 28, 2020 and February 17, 2020. As of February 17, 2020, 420 cases had been cured and discharged from the hospitals. ***Second***, in the isolation wards, the mobile telemedicine device effectively collects, transforms, and evaluates patient health data such as blood pressure, oxygen level, and respiratory rate, and reports them to the care team. This facilitates the avoidance of direct physical contact, thus reducing the risk of exposure to respiratory secretions and preventing the potential transmission of infection to physicians and nurses. ***Third***, the specialist treatment team includes specialists from different clinical disciplines, and can therefore offer comprehensive assessment and treatment. Meanwhile, nurse care managers and social workers can be involved strategically to help patients with pneumonia obtain post-treatment care to avoid coronavirus re-infection. ***Fourth***, the specialist treatment team provides primary care guidance on coronavirus (e.g., clinical criteria for COVID-19 diagnosis, patient transfers, and cleaning process) to all physicians and nurses at 126 connected hospitals via video conference. Substantial efforts are devoted to training physicians and nurses, many of whom are new to the treatment of coronavirus infections.

As we look to the future of epidemic prevention and control, we believe that telemedicine systems have the potential to play a role in addressing emergencies and large-scale outbreaks in high uncertainty settings. As telemedicine has inevitably altered the traditional working relationships within the healthcare network, how to ensure high-quality communication among healthcare practitioners poses a significant challenge. As such, frequent, timely, accurate, and problem-solving focused communication among clinical staffs from hospitals at different levels in the healthcare system is essential to minimize the risk incurred in handling patients with possible COVID-19 infection^3^. However, we have found that high quality of communication is not always maintained during the telemedicine coordination. Therefore, a learning telemedicine system platform for coronavirus care was developed across connected hospitals, serving as the overarching authoritative source for diagnostic decision making and knowledge sharing for treatment. The platform could aggregate COVID-19 patient records across 126 connected hospitals and rapidly expand to enable open collaborations with key stakeholders such as government authorities, research institutions and laboratories. The lessons learned from this crisis can provide insights to guide public health institutions as they implement telemedicine to increase resilience to future epidemic outbreaks.

## Data Availability

The authors confirm that the data supporting the findings of this study are available within the article.

